# Left ventricular concentric remodeling is highly common among veterans previously deployed to Southwest Asia Theater of Military Operations and associated with impaired exercise performance

**DOI:** 10.1101/2024.09.10.24313445

**Authors:** Steven J. Cassady, Thomas J. Abitante, Gregory G. Pappas, Thomas Alexander, Michael J. Falvo, Post-Deployment Cardiopulmonary Evaluation Network

## Abstract

**Background:** Environmental factors, such as exposure to airborne hazards, contribute to cardiac remodeling through a variety of mechanisms including direct cardiotoxicity. Left ventricular concentric remodeling (LVCR) is a pathological process of adaptive myocardial change that may represent a precursor state for systolic and diastolic dysfunction and left ventricular hypertrophy. Given that potentially cardiotoxic airborne hazards, such as those produced by open burn pits, have been found to occur in excess in active military combat zones, deployed veterans may be at increased risk for adverse cardiac remodeling, but this has not been thoroughly investigated.

**Methods:** 139 veterans of Southwest Asia Theater of Military Operations underwent transthoracic echocardiography, cardiopulmonary exercise testing (CPET), and health questionnaires. Two-dimensional echocardiography was used to quantify relative wall thickness (RWT) to classify left ventricular (LV) geometry as normal, concentric/eccentric hypertrophy, or LVCR. Observed rates of LVCR were compared to those reported in the Framingham Heart Study, and CPET results were compared between those with and without LVCR. We examined the association between RWT and select CPET outcomes via an adjusted multivariate regression model.

**Results:** The prevalence of LVCR in the veteran sample (30.2%) was elevated compared to the Framingham Heart Study cohort (6–16%). Demographics and risk factors were similar between veterans with LVCR and normal geometry; however, veterans with LVCR had reduced exercise capacity (V̇O_2,_ 23.7 vs 26.2 ml/kg/min, p<0.05), more inefficient exercise ventilation (VE/V̇CO_2_ nadir: 26.8 vs 25.2, p<0.05), and increased heart rate (HR) reserve (24.7 vs 17.4, p<0.05). RWT was independently associated only with peak HR attained and HR reserve.

**Conclusions:** In our sample of deployed veterans without significant risk factors, the observed rates of LVCR are 2- to 5-fold greater than those reported in a historical civilian cohort. Further, veterans with LVCR also had impaired exercise performance relative to those with normal LV geometry despite otherwise appearing similar. These findings underscore the importance of cardiovascular assessments as part of a dyspnea evaluation for deployed veterans with airborne hazards exposure and raise concerns about their long-term cardiovascular health.

## Background

Left ventricular concentric remodeling (LVCR) is an adaptive response by the myocardium to normalize wall stress brought on by increased cardiac afterload.^1^ This process is reflected by an increase in relative wall thickness (RWT) without a concomitant change in left ventricular mass and carries an elevated risk of development of myocardial dysfunction and systolic and diastolic heart failure, especially amongst hypertensive individuals.^2^ However, altered myocardial geometry, including LVCR, is also found more frequently in normotensive individuals with obesity and/or type 2 diabetes mellitus, suggesting an effect of altered metabolism on the development of these changes. Environmental risk factors such as air pollution, already shown to have a causal relationship with cardiovascular morbidity and mortality, have also been shown to have a variety of cardiotoxic effects, including the development of pathological ventricular remodeling.^3^ Specific mechanisms driving this process may include systemic inflammation, oxidative stress, autonomic nervous system imbalance, and direct cardiotoxicity.^4^

Fine particulate matter (PM_2.5_) air pollution levels are exceedingly high in the military deployment environment of the Southwest Asia Theatre of Military Operations, owing to dust storms, local ambient air pollution, and smoke from open air burn pits.^5^ Service members with longer deployments to military bases with open air burn pits are at demonstrably increased risk of respiratory and cardiovascular conditions, including hypertension.^6^ Given this increased exposure-mediated risk, previously deployed veterans may be at increased risk for cardiac remodeling, which may be further compounded by an increased prevalence of obesity in the veteran population.^7^ To our knowledge, assessment of altered cardiac geometry in deployed veterans has not been systemically investigated. The primary purpose of our study was to investigate the prevalence of left ventricular concentric remodeling (LVCR) in a cohort of treatment-seeking and non-treatment-seeking deployed veterans and to explore the association between altered cardiac geometry and cardiopulmonary function.

## Methods

### Study Participants

Deployed veterans (n = 139) of the Southwest Asia Theater of Military Operations (predominantly post-9/11) who participated in a clinical or research evaluation at our center or affiliated network (2011 – 2024) and had a valid transthoracic echocardiogram comprised our sample. This sample included those who sought treatment (n = 92) and were clinically evaluated at a national Department of Veterans Affairs (VA) specialty clinic (War Related Illness and Injury Study Center)^8^ or national network (Post-Deployment Cardiopulmonary Evaluation Network)^9^ and 47 non-treatment seeking veterans who had volunteered to participate in a research study. All veterans participated in multi-day evaluations that also assessed cardiopulmonary function, evaluated symptoms, and captured medical and military histories. Per our research protocol’s inclusion and exclusion criteria, all 47 non-treatment-seeking volunteers were required to be free of any confirmed or self-reported cardiovascular or respiratory diagnosis at the time of the study. Study procedures were approved and under the oversight of the VA New Jersey Health Care System’s Institutional Review Board and Research and Development Committee (#1577315, 1577279).

All data were abstracted from clinical and research records that included questionnaires, medical and military histories, and in-laboratory assessments. Veterans were classified into normal, overweight, and obese body mass index (BMI) categories – i.e., <25kg/m^2^, ≥ 25kg/m^2^ to <30kg/m^2^, or ≥30 kg/m^2^. The presence or absence of comorbidities of hypertension, diabetes mellitus, and obstructive sleep apnea were confirmed on medical chart review. We also further defined the presence of hypertension based upon a self-report (i.e., those without a local medical record), resting systolic blood pressure ≥140 mmHg or a diastolic blood pressure ≥90 mmHg at the time of visit, or current use of an antihypertensive medication (i.e., angiotensin-converting enzyme inhibitor, angiotensin receptor blocker, beta blockers, calcium channel blocker, or thiazide diuretics).

Health status and symptoms were also assessed via standardized questionnaires including: 1) the Sino-Nasal Outcome Test,^10^ 2) the Modified Medical Research Council dyspnea scale,^11^ 3) the Reflux Disease Questionnaire,^12^ and 4) the Generalized Anxiety Disorder 7 questionnaire.^13^ Also included was a functional activity score, an average of five questions where individuals report the difficulty in performing five basic physical tasks.^14^

### Transthoracic Echocardiography and Left Ventricular Geometry

All participants underwent resting transthoracic echocardiography (TTE) in accordance with published guidelines.^15^ The primary TTE parameters of interest were manually extracted from the clinical report in the electronic health records and included two-dimensional measurements of left ventricular dimensions: left ventricular end-diastolic diameter (LVEDD), posterior wall thickness at end-diastole (PWd), and interventricular septal wall thickness at end-diastole (IVSd), as well as height, weight, and sex to calculate body surface area. We determined the presence and phenotype of left ventricular remodeling by calculating relative wall thickness (RWT) and left ventricular mass index (LVMI). RWT was calculated by the equation (IVSd + PWd)/LVEDD.^16^ A threshold of > 0.42 was used to determine elevated RWT.^17^ We estimated LVMI (g/m^2^) as left ventricular mass calculated by Devereaux’s method^18^ divided by the body surface area. Left ventricular concentric remodeling (LVCR) is defined as a RWT > 0.42 and LVMI ≤ 95 g/m^2^ (women) or ≤ 115 g/m^2^ (men). Concentric and eccentric hypertrophy were defined as a LVMI > 95 g/m^2^ (women) or > 115 g/m^2^ (men) with a RWT > 0.42 or < 0.42, respectively. Rates of LVCR in our population were compared to those reported in the Framingham Heart Study cohort of civilians.^19^ Additional TTE parameters of interest included lateral E/e’ and estimated right ventricular systolic pressure and were also extracted from clinical records.

### Cardiopulmonary Exercise Testing & Pulmonary Function Testing

All veterans underwent a standardized symptom-limited maximal cardiopulmonary exercise test (CPET) using a motor-driven treadmill (Bruce or modified Bruce protocol; n = 28)^20^ or cycle ergometer (25 W/min; n = 84).^21^ All CPETs were assessed for valid effort as defined as a measured peak heart rate (HR) > 85% of age-predicted maximum or a respiratory exchange ratio (RER) > 1.0. Oxygen consumption (V̇O_2_), carbon dioxide production (V̇CO_2_), ventilation (V̇_E_), HR, and O_2_ pulse (V̇O_2_/HR) were obtained breath-by-breath during exercise. Blood pressure was measured every 2 minutes during exercise. Ventilatory efficiency was defined by the V̇_E_/V̇CO_2_ nadir, and the chronotropic response to exercise was calculated per the Wilkoff and Miller method.^22^ Select CPET variables were expressed in absolute values and relative to predicted normal values.^23^ Due to the variation in protocols, only values at peak exercise (average of the last 30 sec.), were utilized for a comparison between the veterans with and without LVCR. On a separate day, complete pulmonary function testing (PFT) was performed in accordance with published standards^24^ using commercially available equipment. PFT variables of interest included those from spirometry (FEV_1_, FVC, and FEV_1_/FVC), body plethysmography (TLC, RV, and RV/TLC), and diffusion capacity (DL_CO_) corrected for hemoglobin, and were expressed both as absolute values and as percent of predicted.^25^

### Statistical Analysis

Rates of LV geometry (normal, concentric/eccentric hypertrophy, or LVCR) were reported for the total cohort, within BMI classes, and amongst veterans with hypertension. Separate reporting by BMI class and hypertension status was performed to allow for qualitative comparison to a published historical sample (Framingham Heart Study).^19^ To account for potential differences related to health-seeking behaviors, we compared sample characteristics that may influence LV geometry and cardiopulmonary outcomes between treatment and non-treatment seeking veteran cohorts. Subsequent analyses (demographics, cardiopulmonary) were performed on the combined group of veterans. Due to the nature of our sample, we anticipated that we would not identify many cases with concentric or eccentric hypertrophy but rather be comprised of either normal LV geometry or LVCR. Therefore, group comparisons were restricted to the combined veteran sample (treatment and non-treatment seeking) with LVCR or normal geometry.

Normality was assessed using the Shapiro-Wilk test. For continuous variables, T-test or Wilcoxon rank-sum test were used as appropriate, with effect sizes reported using Cohen’s d with Hedges’ correction. Chi-squared tests and Cramer’s V were used for categorical variables. Statistical significance was set at *p* < 0.05. Multivariate linear regression models were used to assess the relationship between RWT and select CPET parameters at peak exercise, adjusting for other variables with a known effect on exercise performance (age, BMI, sex, CPET modality), as well as the presence of hypertension. Additional models were used to estimate the adjusted mean differences in CPET outcomes between veterans with and without LVCR. Adjusted associations are presented as beta-coefficients and corresponding 95% confidence intervals and significance. All statistical analyses were performed using R Studio.

## Results

### Sample Description

Baseline characteristics for the treatment (n = 92) and non-treatment (n = 47) seeking veterans are reported in Table 1. Veterans in the treatment group were older, had more comorbidities, had increased lateral E/e’ and right ventricular systolic pressure values, and endorsed more symptoms than non-treatment seeking veterans. Despite similar deployment lengths and time since last deployment, exposure histories differed between groups but were uniformly elevated. A description of our sample and the Framingham Heart Study population used as a historical civilian control comparison of abnormal LV geometry prevalence can be found in the supplemental material (Table S1). Relative to the civilian sample, our combined veteran sample (n = 139) had similar smoking histories but was younger, composed of more men, was more overweight and obese, and had lower rates of hypertension on average. Only one veteran had diabetes mellitus, thereby preventing meaningful comparison to the historical control sample, and the inclusion of this comorbidity was subsequently removed from analysis.

**Table 1:** Demographic characteristics, medical histories, and military histories of study participants, stratified by participant cohort. P values and effect sizes were determined by t-tests (Wilcox as required) and Cohen’s d with hedges correction for continuous variables and with chi-squared and Cramer’s V for categorical variables, respectively.

|  | Non-Treatment (n = 47) |  | Treatment Seeking (n = 92) |  | p value | effect size |
| --- | --- | --- | --- | --- | --- | --- |
|  | n | Mean (SD) | n | Mean (SD) |  |  |
| Age | 47 | 40.36 (7.78) | 92 | 46.27 (9.49) | <0.005 | 0.68 |
| Gender (Female) | 5 | 10.64% | 14 | 15.22% | 0.23 | 0.04 |
| **BMI** | 47 | 28.32 (3.44) | 92 | 31.45 (5.81) | <0.005 | 0.65 |
| Normal | 9 | 19.15% | 8 | 8.70% | <0.01 | 0.26 |
| Overweight | 26 | 55.32% | 36 | 39.13% |  |  |
| Obese | 12 | 25.53% | 48 | 52.17% |  |  |
| **HTN** | 47 |  | 92 |  | <0.001 | 0.29 |
| Yes | 4 | 8.51% | 34 | 36.96% |  |  |
| No | 43 | 91.49% | 58 | 63.04% |  |  |
| **OSA** | 47 |  | 92 |  | <0.001 | 0.61 |
| Yes | 0 | 0.00% | 60 | 65.22% |  |  |
| No | 47 | 100.00% | 32 | 34.78% |  |  |
| **Smoking** | 40 |  | 82 |  | <0.01 | 0.35 |
| Active | 1 | 2.50% | 1 | 1.22% |  |  |
| Former | 7 | 17.50% | 27 | 32.93% |  |  |
| Never | 32 | 80.00% | 54 | 65.85% |  |  |
| Pack Years | 8 | 3.88 (3.44) | 28 | 5.71 (6.15) | 0.42 | 0.32 |
| **Resp Symptoms** |  |  |  |  |  |  |
| Shortness of breath | 47 | 17.02% | 84 | 95.24% | <0.001 | 0.78 |
| Sputum | 47 | 23.40% | 84 | 71.43% | <0.001 | 0.45 |
| Cough | 47 | 8.51% | 85 | 68.24% | <0.001 | 0.56 |
| **Questionnaires** |  |  |  |  |  |  |
| SNOT22<sup>1</sup> | 47 | 20.17 (16.53) | 83 | 41.33 (20.92) | <0.005 | 1.12 |
| MMRC<sup>2</sup> | 36 | 0.47 (0.61) | 81 | 1.48 (0.91) | <0.005 | 1.30 |
| Functional Score<sup>3</sup> | 47 | 1.57 (0.68) | 83 | 2.59 (0.90) | <0.005 | 1.28 |
| RDQ<sup>4</sup> | 47 | 1.47 (2.57) | 85 | 6.18 (5.16) | <0.005 | 1.15 |
| GAD7<sup>5</sup> | 47 | 4.96 (6.11) | 68 | 8.52 (6.02) | <0.005 | 0.58 |
| **Military History** |  |  |  |  |  |  |
| Total Deploy Length (Days) | 47 | 535.57 (375.38) | 88 | 475.63 (376.00) | 0.18 | 0.16 |
| Months Since Last Deploy | 47 | 141.32 (50.10) | 88 | 161.36 (76.81) | 0.22 | 0.31 |
| Months Since First Exposure | 47 | 173.61 (42.33) | 88 | 219.41 (89.45) | <0.005 | 0.65 |
| **Exposures** |  |  |  |  |  |  |
| Blast | 47 | 68.09% | 90 | 40.00% | <0.005 | 0.30 |
| Dust storms | 47 | 85.11% | 81 | 96.30% | <0.05 | 0.27 |
| Burn pits | 47 | 80.85% | 81 | 61.73% | <0.005 | 0.32 |
| Vehicle Exhaust | 47 | 72.34% | 81 | 83.95% | 0.18 | 0.12 |
| **Echo** |  |  |  |  |  |  |
| RWT | 47 | 0.36 (0.10) | 92 | 0.40 (0.09) | <0.05 | 0.43 |
| LVMI (g/m<sup>2</sup>) | 47 | 69.57 (20.29) | 92 | 71.63 (19.18) | 0.67 | 0.10 |
| Lateral E/e' | 47 | 5.80 (1.52) | 62 | 4.98 (2.84) | <0.05 | 0.52 |
| RVSP (mmHg) | 41 | 20.06 (6.06) | 25 | 25.01 (7.09) | <0.05 | 0.75 |

1. SinoNasal Outcome Test: Score is sum of 22 questions concerning upper respiratory & sinus symptoms. Answers range from 0 (no symptom) to 5 (severe symptoms)
2. Modified Medical Research Council Dyspnea Scale: 1 (only breathless during strenuous exercise) to 5 (breathless during basic living activity)
3. Mean score of 5 questions rating the difficulty of basic daily activities (e.g. walk 1 mile, walk up steep hill, 12 stairs). Answers range from 0 (no difficulty) to 5 (incapable)
4. Reflux Disease Questionnaire: Total score is mean of 3 different survey scores concerning the frequency and intensity of heartburn, regurgitation, or dyspeptic complaints respectively. Each sub survey is the sum of 4 questions. Answers range from 0 (no symptom) to 5 (severe symptoms)
5. Generalized Anxiety Disorder Scale 7: Score is sum of 7 questions concerning the frequency of anxiety symptoms. Answers range from 0 (never) to 3 (nearly every day)

### Transthoracic Echocardiography and Left Ventricular Geometry

Abnormal LV geometry was present in 33.8% (47 of 139) of our sample, characterized predominantly by LVCR (42 of 47). Rates of LVCR were similar between non-treatment (27.7%, 13 of 47) and treatment-seeking veterans (31.5%, 29 of 92). Figure 1 illustrates LVCR prevalence for the combined veteran sample as a function of BMI class and hypertension status along with comparison to the Framingham cohort. In our sample, LVCR prevalence was higher among veterans who were overweight or obese (31.6% – 33.9%) relative to those with normal weight (11.8%) but was similar among those with (31.6%) or without (29.7%) hypertension. When compared to the Framingham Cohort, rates of LVCR were higher among veterans for all comparisons with mean absolute differences ranging from +3.1% to +20.2%. A complete description of the LV geometry rates in our sample can be found in the supplemental material (Table S2).

**Figure 1.**
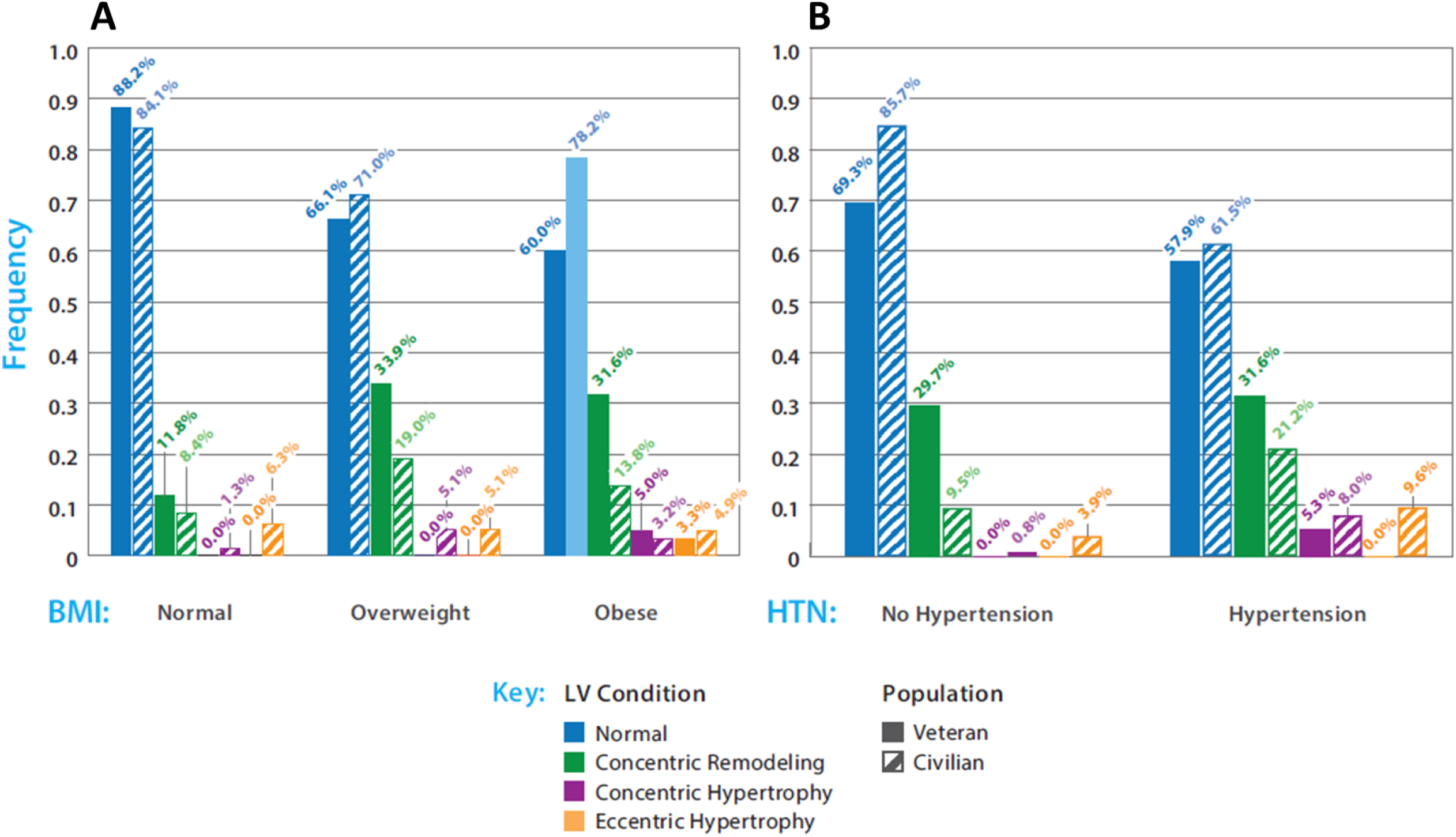
Frequency of LV concentric remodeling in our deployed veteran sample (n = 139) compared to a historical civilian population as a function of A) body mass index (BMI) and B) presence of hypertension (HTN). Historical civilian population rates were extrapolated from Von Jenison et al 2020 (n = 5,432), The civilian sample extrapolation excluded subjects with diabetes.

We restricted our analysis to the 134 veterans with normal LV geometry or LVCR and excluded those with concentric (n = 3) and eccentric (n = 2) hypertrophy. Demographics are reported in Table 2. Ten veterans in the treatment-seeking group were described on physician read of their echocardiogram as having at least mild left ventricular hypertrophy, and two were read as having moderate or severe reductions in left ventricular ejection fraction. Veterans in the LVCR group were slightly older and had slightly higher rates of overweight and obesity, though BMI was not significantly different between the groups. Self-reported airborne hazards exposure from four domains (open burn pits, vehicle exhaust, dust storms, and blast exposures) were similar; however, those with LVCR had a greater total deployment length (Table 2). LVMI, lateral E/e’ and estimated right ventricular systolic pressure were similar between groups.

**Table 2:**
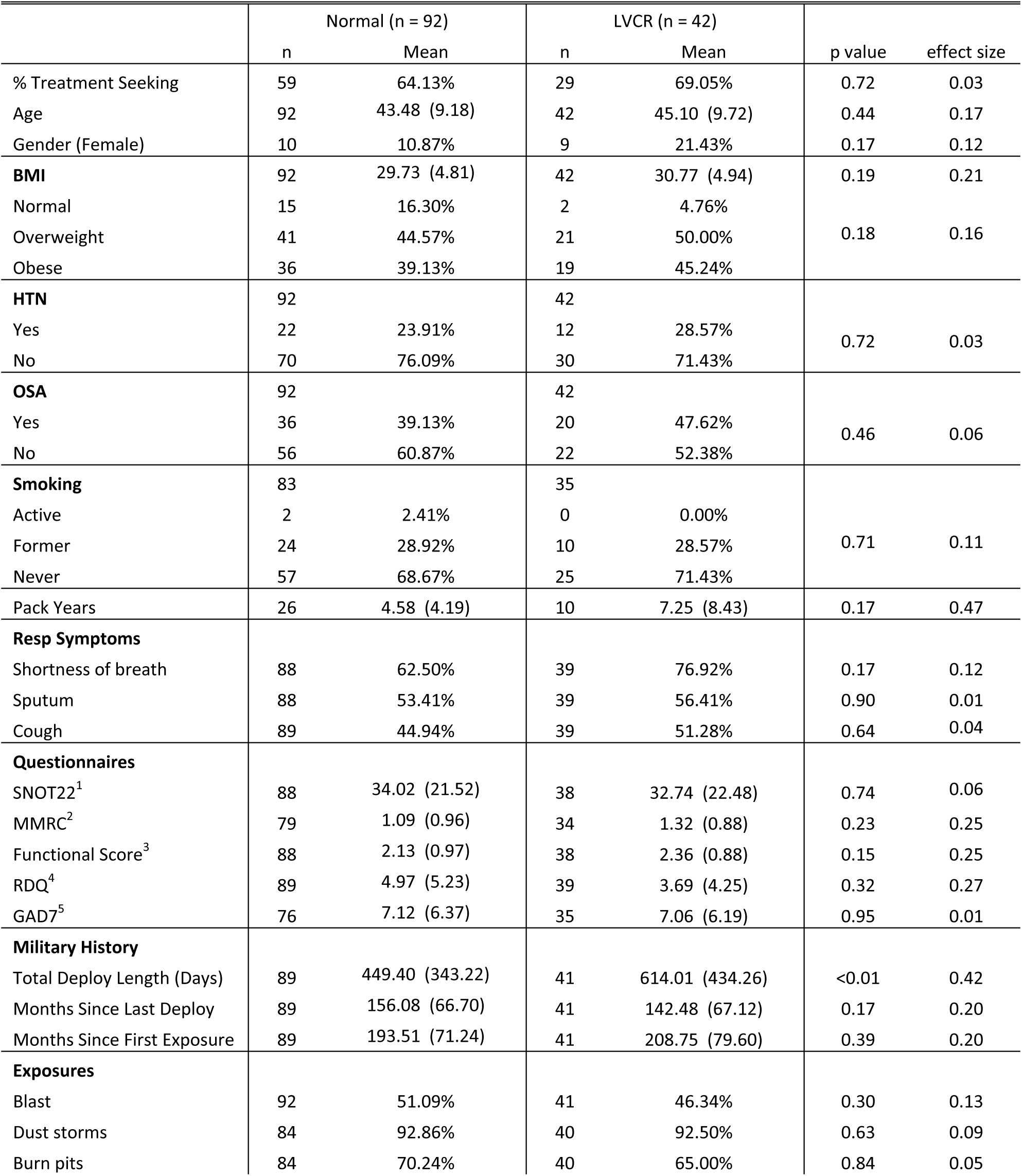
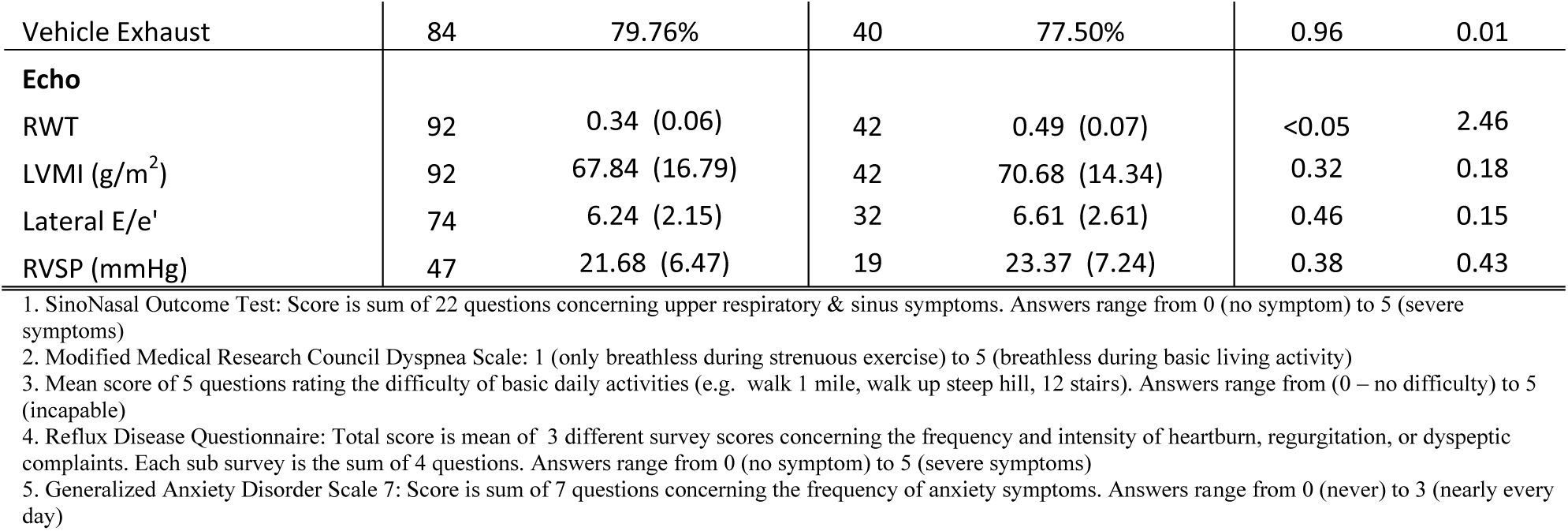
Demographic characteristics, medical histories, and military histories of study participants, stratified by left ventricular geometry, as determined by a resting transthoracic echocardiogram. Participants with either concentric or eccentric hypertrophy (n = 5) were excluded. p values and effect sizes were determined by t-tests (Wilcox as required) and Cohen’s d with hedges correction for continuous variables and with chi-squared and Cramer’s V for categorical variables, respectively.

### Cardiopulmonary Exercise Testing & Pulmonary Function Testing

CPET data were available for 112 of 134 veterans with 105 achieving maximal effort (Figure 2). Unadjusted comparison of CPET performance between the normal and LVCR groups is shown in Table 3. Exercise capacity (V̇O_2_, ml/kg/min) was reduced on average (*d* = 0.41) and ventilation was less efficient (V_E_/V̇CO_2_ nadir; *d* = 0.48) among veterans with LVCR in comparison to those with normal LV geometry. The heart rate response to exercise was also blunted in those with LVCR relative to normal LV geometry as evidenced by a reduced chronotropic index, reduced peak predicted heart rate, and increased heart reserve with small-to-moderate effect sizes (*d* = 0.32 to 0.55).

**Figure 2:**
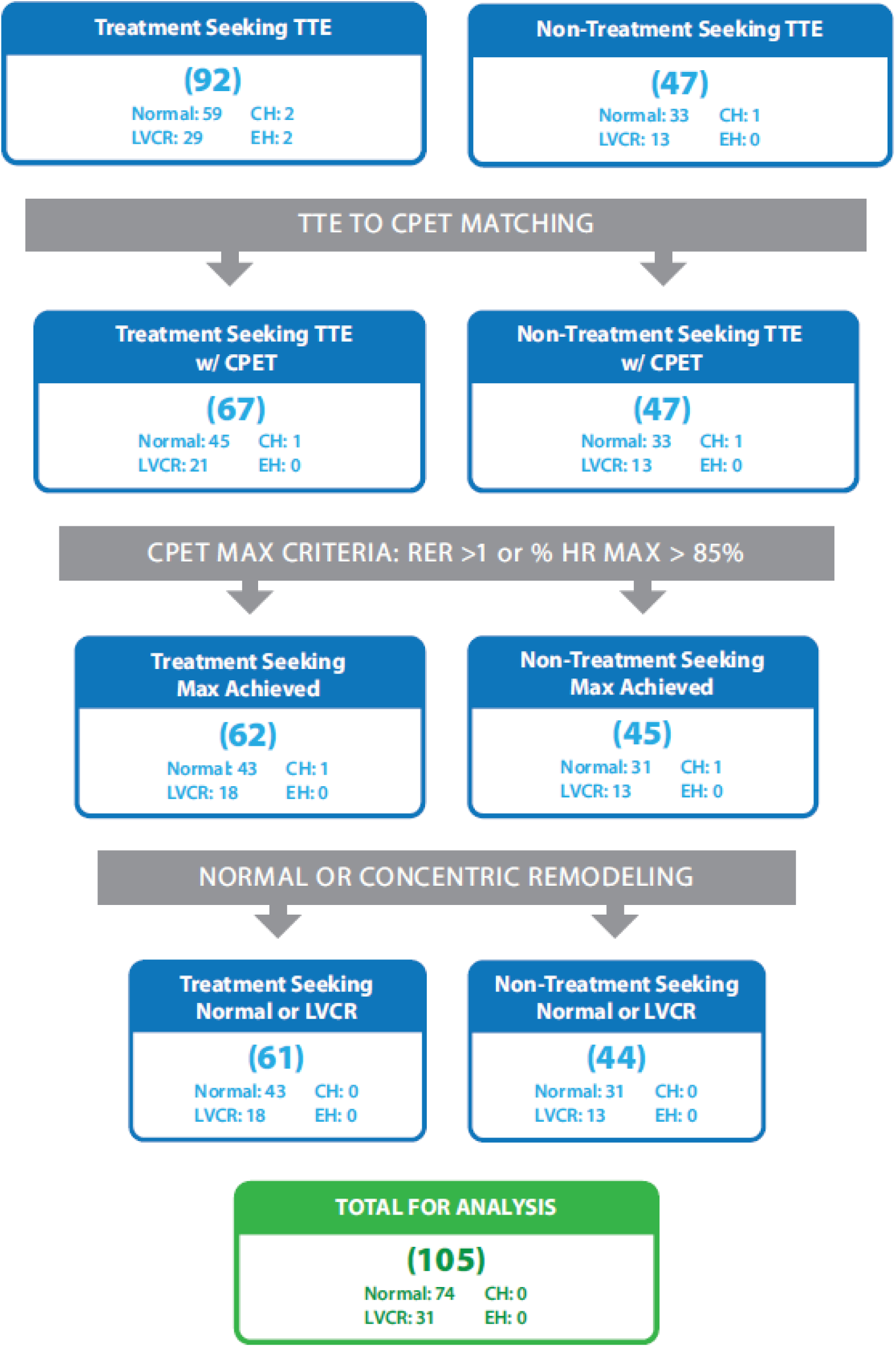
Flow diagram of the CPET filtering workflow, from the initial complete participant sample to those included in the CPET analysis, separated by participant cohort (Non-Treatment vs Treatment). TTE: Transthoracic Echocardiogram, LVCR: Left Ventricular Concentric Remodeling, CH: Concentric Hypertrophy, EH: Eccentric Hypertrophy, RER: Respiratory Exchange Ratio.

**Table 3:** Summary statistics of CPET parameters for veterans with and without LV Concentric Remodeling (LVCR). CPET parameters denote value at peak exercise unless otherwise stated. Δ Pressures are the difference from rest to peak.

|  | Normal (n = 74) |  |  | LVCR (n=31) |  |  | p value | d value |
| --- | --- | --- | --- | --- | --- | --- | --- | --- |
|  | n | Mean | SD | n | Mean | SD |  |  |
| $\dot{V}O_2$ max (ml/min) | 74 | 2360.43 | 629.11 | 31 | 2210.26 | 645.91 | 0.28 | 0.23 |
| $\dot{V}O_2$/kg max (ml/kg/min) | 74 | 26.16 | 6.69 | 31 | 23.68 | 5.31 | <0.05 | 0.41 |
| $\dot{V}O_2$/kg max pp | 73 | 80.10 | 16.72 | 31 | 78.05 | 15.65 | 0.55 | 0.13 |
| $\dot{V}E/\dot{V}CO_2$ Nadir<sup>1</sup> | 75<sup>a</sup> | 25.15 | 3.17 | 31 | 26.80 | 3.72 | <0.05 | 0.48 |
| O<sub>2</sub> Pulse (ml/beat) | 73 | 14.75 | 3.90 | 31 | 14.58 | 3.79 | 0.42 | 0.04 |
| Chronotropic Response | 70 | 1.16 | 0.32 | 31 | 1.06 | 0.31 | 0.10 | 0.32 |
| HR max pp | 73 | 91.08 | 6.87 | 31 | 86.50 | 9.50 | <0.05 | 0.55 |
| HR Reserve @ Peak (bpm) | 73 | 17.37 | 11.88 | 31 | 24.73 | 15.69 | <0.05 | 0.53 |
| $\Delta$ Pulse Pressure (mmHg) | 67 | 29.40 | 34.10 | 28 | 26.21 | 32.69 | 0.67 | 0.09 |
| Diastolic @ peak (mmHg) | 73 | 86.05 | 17.57 | 31 | 87.68 | 16.11 | 0.71 | 0.10 |
| $\Delta$ Diastolic (mmHg) | 68 | 5.28 | 13.94 | 29 | 7.55 | 12.85 | 0.40 | 0.17 |
| Power @ Max (Watts) | 57 | 186.84 | 41.53 | 25 | 175.60 | 46.00 | 0.30 | 0.25 |

1. $\dot{V}E/\dot{V}CO_2$ Nadir analysis performed with all CPETS, including those without an achieved max (n = 112)

The results of the multivariate linear regression model are depicted in Figure 3. In the fully adjusted model, only achieved age-predicted maximal HR and HR reserve at peak exercise were significantly associated with RWT. Additionally, an adjusted means analysis comparing the veterans with and without LVCR found a significant difference only between achieved age-predicted maximal HR and HR reserve at peak when correcting for other parameters (Table 4).

**Figure 3:**
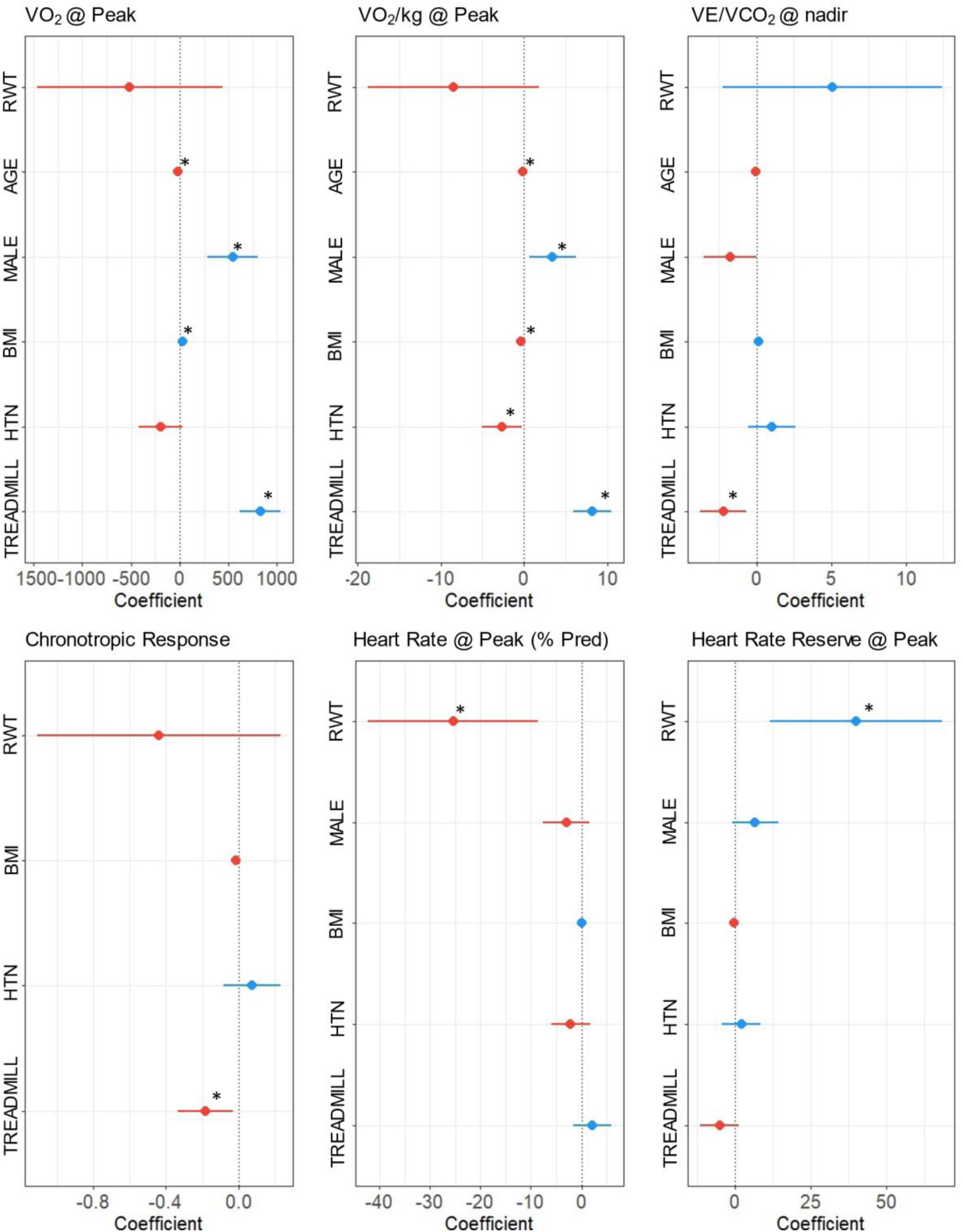
Multivariate linear regression model coefficients (95% CI) for select CPET parameters, * p <0.05

**Table 4:** Adjusted mean differences of CPET parameters between veterans with and without LV concentric remodeling. Adjusted for parameters with known effects of CPET performance (Age, Sex, BMI) as well as the presence of hypertension.

|  | adj mean diff | p value |
| --- | --- | --- |
| $\dot{V}O_2$ max (ml/min) | 44.97 | 0.64 |
| $\dot{V}O_2$/kg max (ml/kg/min) | 1.07 | 0.30 |
| $\dot{V}O_2$/kg max pp | 1.27 | 0.70 |
| $\dot{V}E/\dot{V}CO_2$ Nadir | -1.33 | 0.06 |
| O<sub>2</sub> Pulse (ml/beat) | -0.46 | 0.45 |
| Chronotropic Response | 0.12 | 0.08 |
| HR max pp | 4.71 | <0.05 |
| HR Reserve @ Peak (bpm) | -7.89 | <0.05 |
| $\Delta$ Pulse Pressure (mmHg) | -0.39 | 0.96 |
| Diastolic @ peak (mmHg) | 1.02 | 0.76 |
| $\Delta$ Diastolic (mmHg) | -1.04 | 0.73 |
| Power @ Max (Watts) | 4.67 | 0.60 |

PFT data were available for 129 of 134 veterans. Spirometry, lung volume and diffusion were similar between veterans with LVCR and normal LV geometry (Table 5). There were no significant differences in any of the percent predicted lung volumes or ratios. Finally, diffusion capacity for carbon monoxide (D_LCO_) did not differ between groups.

**Table 5:** Summary statistics of PFT parameters for veterans with and without LV Concentric Remodeling (LVCR).

|  | Normal (n = 90) |  |  | LVCR (n=39) |  |  | p value | d value |
| --- | --- | --- | --- | --- | --- | --- | --- | --- |
|  | n | Mean | SD | n | Mean | SD |  |  |
| FEV1 | 90 | 3.75 | 0.87 | 39 | 3.46 | 0.90 | 0.09 | 0.33 |
| FEV1 pp | 88 | 96.98 | 16.84 | 39 | 92.38 | 19.82 | 0.21 | 0.25 |
| FVC (L) | 90 | 4.78 | 1.02 | 39 | 4.33 | 1.13 | <0.05 | 0.42 |
| FVC pp | 88 | 99.33 | 14.38 | 39 | 93.16 | 18.66 | 0.07 | 0.37 |
| FEV1/FVC | 90 | 78.38 | 7.22 | 39 | 79.85 | 5.38 | 0.40 | 0.23 |
| FEV1/FVC pp | 88 | 97.09 | 8.69 | 39 | 99.24 | 7.81 | 0.27 | 0.26 |
| TLC (L) | 89 | 6.56 | 1.38 | 38 | 6.00 | 1.16 | <0.05 | 0.44 |
| TLC pp | 87 | 94.43 | 15.28 | 38 | 89.44 | 13.99 | 0.08 | 0.34 |
| RV (L) | 89 | 1.69 | 0.87 | 38 | 1.59 | 0.61 | 0.45 | 0.14 |
| RV pp | 87 | 96.99 | 44.90 | 38 | 93.11 | 30.94 | 0.58 | 0.10 |
| RV/TLC | 89 | 25.03 | 10.29 | 38 | 26.98 | 10.00 | 0.32 | 0.19 |
| RV/TLV pp | 87 | 101.09 | 38.31 | 38 | 105.76 | 32.79 | 0.49 | 0.13 |
| DLCO pp | 87 | 95.23 | 14.56 | 38 | 93.40 | 16.08 | 0.55 | 0.12 |

## Discussion

In our sample of veterans deployed to the Southwest Asia Theater of Military Operations, approximately 1/3^rd^ had evidence of left ventricular (LV) remodeling, specifically LV concentric remodeling (LVCR), as assessed by 2D echocardiography. Further, we found that exercise capacity was reduced among veterans with LVCR and that an index of LV geometry – i.e., relative wall thickness (RWT) – independently predicts maximal heart rate achieved during CPET (Figure 3). The rates of remodeling found are markedly higher than those reported in a large civilian cohort with known risk factors (i.e., hypertension and diabetes mellitus) for cardiac remodeling (Figure 1). That LVCR was present amongst both treatment and non-treatment seeking cohorts also suggests that this finding may be common among previously deployed individuals. Our findings of LV remodeling and its association with exercise performance are concerning and highlight the importance of considering non-pulmonary contributions to exertional dyspnea in deployed veterans with military environmental exposures.

LV remodeling is a pathological, adaptive myocardial process that includes cardiomyocyte hypertrophy, apoptosis, and excess collagen deposition leading to interstitial myocardial fibrosis.^26^ This process has been described as a generalized adaptive response to increased wall stress, mediated by alternations in the neuro-endocrine axis, particularly the renin-angiotensin-aldosterone system, sympathetic nervous system, systemic inflammation, and oxidative stress, ultimately creating a thicker LV wall and a resultant decrease in LV chamber size. These physiologic changes are thought to mirror subclinical compensatory changes that are typically unaccompanied by symptoms, allowing for preservation of LV systolic function. LV remodeling can be assessed with several different imaging techniques, of which echocardiography remains the most widely utilized. The three patterns of LV remodeling determined by two-dimensional echocardiography – concentric remodeling, concentric hypertrophy, and eccentric hypertrophy -were explored initially in the VALIANT study and their definitions have become universally accepted.^27^ Regardless of pattern, this remodeling is thought to be an intermediate step towards the development of heart failure, with possible far-reaching systemic consequences, including associations with pulmonary airflow obstruction, chronic renal disease, anemia, and systemic inflammation.^28^

In this study, veterans with LVCR were found to have reduced exercise capacity (V̇O_2_ peak) compared to those without this remodeling. Studies of patients with LV remodeling suggest that V̇O_2_ peak is linked to a reduction in LV end-diastolic volume, as the LV chamber decreases in size as the remodeled walls thicken.^29^ Though increased wall thickness and eccentric LV hypertrophy are adaptations that can also be seen in highly conditioned athletes, these changes do not correlate with systolic or diastolic dysfunction and in fact result in higher peak V̇O_2_ values, with end-diastolic volume of the left ventricle shown to increase rather than decrease.^30^ Additionally, the LV remodeling group had a higher V̇E/V̇CO_2_ nadir than the normal group; this value is commonly used to determine the presence of ventilatory inefficiency, with higher values correlated with worsened outcomes in heart failure patients.^31^ Relative chronotropic incompetence was suggested in the LV remodeling group, with higher heart rate reserve and lower heart rates based on percent predicted values at peak exercise. These findings suggest a subtler manifestation of the chronotropic incompetence described in patients with increased LV mass and cavity size in the Framingham Study cohort, in which LV mass above the 90^th^ percentile was associated with failure to achieve target predicted heart rates.^32^ Finally, and unexpectedly given the known association between LV remodeling and obstructive lung disease and the nature of the inhaled toxins implicated, there was no suggestion of an increase in spirometric obstruction. In fact, percent-predicted values for forced vital capacity (FVC) and total lung capacity (TLC) were reduced in the LVCR group, suggesting a tendency towards restriction rather than obstruction that may reflect the slightly elevated BMI in the LVCR group.

The possible underlying factors leading to higher rates of pathological LV concentric remodeling amongst deployed veterans are numerous. Cardiac remodeling is influenced by the aging process and by the presence of systemic hypertension, sleep apnea, diabetes mellitus, and obesity,^28^ all of which, save diabetes mellitus, were relatively frequent comorbidities amongst our veteran cohort. Notably, rates of obesity were higher in the veterans who sought treatment for unexplained dyspnea. However, even with these comorbidities, prevalence rates for remodeling were higher than would have been expected, and both the treatment-seeking and non-treatment-seeking cohorts were relatively young to develop a process typically associated with “normal aging.” Aside from comorbidities, common to the entire cohort is a history of frequent exposure to airborne hazards during overseas military service. Air pollution has been clearly demonstrated as a causal factor behind the development of cardiovascular disease in large civilian cohorts.^3,33^ Exposure to a wide range of inhaled pollutants is a frequent and pervasive occurrence for veterans deployed to warzones in the Southeast Asia Theater of Military Operations, including local ambient air pollution, engine exhaust, dust storms, and, perhaps most prominently, smoke generated from burn pits used for waste disposal.^34^

Burn pits are used to dispose of a variety of wastes, including chemicals, medical and human waste, metals, rubber, wood, food waste, and petroleum and plastic products.^35^ Specific toxins identified from the burning of these wastes include particulate matter (PM), airborne metal particles, benzene, polychlorinated dibenzo-p-dioxins and dibenzo-p-furans (PCDDs/Fs), polycyclic aromatic hydrocarbons (PAHs), and volatile organic compounds (VOCs).^36^ There exists substantial evidence to support the presence of cardiotoxicity from each of these categories of pollutants *in vitro* and some evidence from population-based studies to suggest cardiotoxic effects in humans. Benzene exposure raises circulating levels of angiogenic cells,^37^ which may indicate stress to the endothelium, and increases inflammation and cell-cell adhesion in the myocardium of a mouse model of hypertension-induced heart failure.^38,39^ Dioxin-like compounds, a specific class of PAH, have been shown as far back as 2001 to produce direct cardiovascular toxicity in animal models,^40–42^ with chronic exposure producing increases in systemic blood pressure, circulating cholesterol and triglycerides, and resulting in chronic arteritis, myocardial hypertrophy, and cardiomyopathy.^43^ These effects were found to be linked to increases in inflammation, oxidative stress, and possible mitochondrial dysfunction.^44^ Finally, smoke from combustion of plastic waste has been shown to produce higher amounts of systemic inflammation and lung injury than non-plastic-containing samples.^45^ These cardiotoxic effects were recently examined more directly in humans with the findings that microplastic and nanoplastic content in carotid artery plaques conferred a higher risk of myocardial infarction, stroke, or death.^46^ Larger studies in humans will be critical to establish the connection between inhalation of the toxic compounds emanated by burn pits and the development of cardiotoxic effects, and to determine the factors underlying differential development of cardiotoxicity between participants with similar exposure patterns.

Beyond the effects of specific pollutants, the effects of both the duration and intensity of air pollutant exposure on cardiovascular disease have been the subject of intense investigation. Both long duration, low intensity exposure, such as that encountered in urban and traffic associated pollution,^47^ or short duration, high intensity exposure, like that experienced by disaster relief workers, have been shown to result in adverse concentric remodeling.^48^ In older individuals, short duration, low intensity exposure can also greatly increase the risk of developing cardiovascular disease.^49^ Military veterans deployed to Southwest Asia Theater of Military Operations represent a unique population exposed to a wide range of air pollutants at intense levels; in these countries, reported PM_2.5_ levels have exceeded 10,000 µg/m^3,50^ far above the 25 µg/m^3^ level set as the allowable daily exposure by the World Health Organization. This high level of PM is derived from both geologic sources (e.g., dust storms) and anthropogenic sources, including industrial pollution, civilian and military combustion, and burn pits.^35,51^ Due to the average total deployment length of 7.3 to 17.8 months (based on service and component)^52^, veterans experience a unique pattern of exposure to inhaled pollutants characterized by moderate duration and high intensity. Any long-term physiological effects of the exposures may be exacerbated by the unique characteristics of combat experience that can make an individual more susceptible to the effects of air pollution, such as high stress,^53^ high physical demands^54^, and the requirements of certain military occupational specialties,^55^ all of which may influence the development of systemic hypertension.^56,57^ Cardiovascular outcomes and rates of concentric remodeling from such moderate duration, high intensity exposures up to this point have not been well-demonstrated.

This study has several notable strengths and limitations. A strength of this study is the inclusion of two separate deployed veteran cohorts – i.e., treatment and non-treatment-seeking – who were evaluated nationally across five separate VA medical centers. Several limitations should be acknowledged regarding available data. Although efforts were made to maximize similar data across cohorts, exercise and echocardiographic data were incomplete or missing for some veterans, and only resting echocardiographic measurements were available, which may limit the potential to evaluate diastolic dysfunction more likely to emerge only with provocation. Available rates of LV remodeling in the general population are sparse, and comparisons of our veteran cohort with a large civilian cohort should be interpreted with caution, as our veteran cohort was younger and predominantly male. However, given the strong association of LV remodeling with increasing age,^58^ this may serve to strengthen our conclusions.

## Conclusions

In this study of veterans formerly deployed to predominantly post-9/11 conflicts in the Southwest Asia Theater of Military Operations, rates of LV remodeling were markedly higher than expected rates established by a large population-based study, the Framingham Heart Study. This elevated prevalence was found to be the case both for veterans who presented to specialized evaluation centers for unexplained dyspnea and those who did not. The presence of LV remodeling was associated with multiple abnormalities during maximal cardiopulmonary exercise testing, including a significant reduction in exercise capacity, ventilatory efficiency, and heart rate response to exercise. The rates of LV remodeling may have been influenced to a modest degree by relatively high rates of obesity but does not fully explain our findings. More research is required to determine if specific airborne hazard exposures represent a likely mediator for development of this pathophysiological change to the myocardium.

## Data Availability

All data were collected at the VA and the signed subject consent forms did not make provision for making individual data records publicly available, even in de-identified form. However the authors can provide the ?metadata? ? i.e. the numerical (aggregated data) results used to generate the figures. Requests for access can be sent to: Joselyn McLaughlin, PhD. Research Compliance Officer Director's Office East Orange Campus of the VA New Jersey Healthcare System 385 Tremont Ave East Orange, NJ 07018 PH: 973-676-1000 x2035

## Acknowledgements

The authors thank the Veterans who volunteered for this study or who were evaluated within the VA’s Post-Deployment Cardiopulmonary Evaluation Network (PDCEN). We would also like to thank the following colleagues who acquired, contributed, and/or organized data: PDCEN Site Directors (Drs. Mehrdad Arjomandi, Danielle Glick, Silpa Krefft, John Osterholzer, Bradley Richmond, Anays Sotolongo) and their respective teams; Courtney Eberhardt and the PDCEN coordinators; Nicole Piskura and the Cardiorespiratory Physiology Laboratory.

## Supplemental Material

1. Subgroup Comparisons (Non-Treatment vs Treatment vs Combined vs Framingham Summary)
2. Subgroup LV geometry Rates (Non-Treatment vs Treatment vs Combined)

## References

1. Rosen BD, Edvardsen T, Lai S, Castillo E, Pan L, Jerosch-Herold M, Sinha S, Kronmal R, Arnett D, Crouse JR, et al. Left Ventricular Concentric Remodeling Is Associated With Decreased Global and Regional Systolic Function: The Multi-Ethnic Study of Atherosclerosis. Circulation. 2005;112:984–991.

2. Nauta JF, Hummel YM, Tromp J, Ouwerkerk W, van der Meer P, Jin X, Lam CSP, Bax JJ, Metra M, Samani NJ, et al. Concentric vs. eccentric remodelling in heart failure with reduced ejection fraction: clinical characteristics, pathophysiology and response to treatment. Eur. J. Heart Fail. 2020;22:1147–1155.

3. Brook RD, Rajagopalan S, Pope CA, Brook JR, Bhatnagar A, Diez-Roux AV, Holguin F, Hong Y, Luepker RV, Mittleman MA, et al. Particulate Matter Air Pollution and Cardiovascular Disease: An Update to the Scientific Statement From the American Heart Association. Circulation. 2010;121:2331–2378.

4. Liu Y, Goodson JM, Zhang B, Chin MT. Air pollution and adverse cardiac remodeling: clinical effects and basic mechanisms. Front. Physiol. [Internet]. 2015 [cited 2024 Jan 3];6. Available from: https://www.frontiersin.org/articles/10.3389/fphys.2015.00162

5. Engelbrecht JP, McDonald EV, Gillies JA, Jayanty RKM, Casuccio G, Gertler AW. Characterizing Mineral Dusts and Other Aerosols from the Middle East—Part 1: Ambient Sampling. Inhal. Toxicol. 2009;21:297–326.

6. Savitz DA, Woskie SR, Bello A, Gaither R, Gasper J, Jiang L, Rennix C, Wellenius GA, Trivedi AN. Deployment to Military Bases With Open Burn Pits and Respiratory and Cardiovascular Disease. *JAMA Netw*. Open. 2024;7:e247629.

7. Breland JY, Phibbs CS, Hoggatt KJ, Washington DL, Lee J, Haskell S, Uchendu US, Saechao FS, Zephyrin LC, Frayne SM. The Obesity Epidemic in the Veterans Health Administration: Prevalence Among Key Populations of Women and Men Veterans. J. Gen. Intern. Med. 2017;32:11–17.

8. Lange G, McAndrew L, Ashford JW, Reinhard M, Peterson M, Helmer DA. War Related Illness and Injury Study Center (WRIISC): A Multidisciplinary Translational Approach to the Care of Veterans With Chronic Multisymptom Illness. Mil. Med. 2013;178:705–707.

9. Davis CW, Rabin AS, Jani N, Osterholzer JJ, Krefft S, Hines SE, Arjomandi M, Robertson MW, Sotolongo AM, Falvo MJ. Postdeployment Respiratory Health: The Roles of the Airborne Hazards and Open Burn Pit Registry and the Post-Deployment Cardiopulmonary Evaluation Network. Fed. Pract. 2022;39:337–343.

10. Hopkins C, Gillett S, Slack R, Lund V j., Browne J p. Psychometric validity of the 22-item Sinonasal Outcome Test. Clin. Otolaryngol. 2009;34:447–454.

11. Fletcher CM, Elmes PC, Fairbairn AS, Wood CH. Significance of Respiratory Symptoms and the Diagnosis of Chronic Bronchitis in a Working Population. Br. Med. J. 1959;2:257–266.

12. Shaw MJ, Talley NJ, Beebe TJ, Rockwood T, Carlsson R, Adlis S, Fendrick AM, Jones R, Dent J, Bytzer P. Initial validation of a diagnostic questionnaire for gastroesophageal reflux disease. Am. J. Gastroenterol. 2001;96:52–57.

13. Spitzer RL, Kroenke K, Williams JBW, Löwe B. A Brief Measure for Assessing Generalized Anxiety Disorder: The GAD-7. Arch. Intern. Med. 2006;166:1092.

14. National Academies of Sciences E and Medicine. Assessment of the Department of Veterans Affairs Airborne Hazards and Open Burn Pit Registry [Internet]. Washington, DC: The National Academies Press; 2017. Available from: https://nap.nationalacademies.org/catalog/23677/assessment-of-the-department-of-veterans-affairs-airborne-hazards-and-open-burn-pit-registry

15. Mitchell C, Rahko PS, Blauwet LA, Canaday B, Finstuen JA, Foster MC, Horton K, Ogunyankin KO, Palma RA, Velazquez EJ. Guidelines for Performing a Comprehensive Transthoracic Echocardiographic Examination in Adults: Recommendations from the American Society of Echocardiography. J. Am. Soc. Echocardiogr. 2019;32:1–64.

16. Yamaguchi S, Shimabukuro M, Abe M, Arakaki T, Arasaki O, Ueda S. Comparison of the prognostic values of three calculation methods for echocardiographic relative wall thickness in acute decompensated heart failure. Cardiovasc. Ultrasound. 2019;17:30.

17. Lang RM, Badano LP, Mor-Avi V, Afilalo J, Armstrong A, Ernande L, Flachskampf FA, Foster E, Goldstein SA, Kuznetsova T, et al. Recommendations for Cardiac Chamber Quantification by Echocardiography in Adults: An Update from the American Society of Echocardiography and the European Association of Cardiovascular Imaging. J. Am. Soc. Echocardiogr. 2015;28:1–39.e14.

18. Devereux RB. Detection of left ventricular hypertrophy by M-mode echocardiography. Anatomic validation, standardization, and comparison to other methods. Hypertens. Dallas Tex 1979. 1987;9:II19-26.

19. von Jeinsen B, Vasan RS, McManus DD, Mitchell GF, Cheng S, Xanthakis V. Joint influences of obesity, diabetes, and hypertension on indices of ventricular remodeling: Findings from the community-based Framingham Heart Study. PLoS ONE. 2020;15:e0243199.

20. Okin PM, Ameisen O, Kligfield P. A modified treadmill exercise protocol for computer-assisted analysis of the ST segment/heart rate slope: Methods and reproducibility. J. Electrocardiol. 1986;19:311–318.

21. ATS/ACCP Statement on Cardiopulmonary Exercise Testing. Am. J. Respir. Crit. Care Med. 2003;167:211–277.

22. Wilkoff BL, Miller RE. Exercise testing for chronotropic assessment. Cardiol. Clin. 1992;10:705– 717.

23. De Souza E Silva CG, Kaminsky LA, Arena R, Christle JW, Araújo CGS, Lima RM, Ashley EA, Myers J. A reference equation for maximal aerobic power for treadmill and cycle ergometer exercise testing: Analysis from the FRIEND registry. Eur. J. Prev. Cardiol. 2018;25:742–750.

24. Stanojevic S, Kaminsky DA, Miller MR, Thompson B, Aliverti A, Barjaktarevic I, Cooper BG, Culver B, Derom E, Hall GL, et al. ERS/ATS technical standard on interpretive strategies for routine lung function tests. Eur. Respir. J. 2022;60:2101499.

25. Quanjer PH, Stanojevic S, Cole TJ, Baur X, Hall GL, Culver BH, Enright PL, Hankinson JL, Ip MSM, Zheng J, et al. Multi-ethnic reference values for spirometry for the 3–95-yr age range: the global lung function 2012 equations. Eur. Respir. J. 2012;40:1324–1343.

26. Konstam MA, Kramer DG, Patel AR, Maron MS, Udelson JE. Left Ventricular Remodeling in Heart Failure. JACC Cardiovasc. Imaging. 2011;4:98–108.

27. Velazquez EJ, Pfeffer MA, McMurray JV, Maggioni AP, Rouleau J-L, Van de Werf F, Kober L, White HD, Swedberg K, Leimberger JD, et al. VALsartan In Acute myocardial iNfarcTion (VALIANT) trial: baseline characteristics in context. Eur. J. Heart Fail. 2003;5:537–544.

28. Cheng S, Vasan RS. Advances in the Epidemiology of Heart Failure and Left Ventricular Remodeling. Circulation. 2011;124:e516–e519.

29. Letnes JM, Nes BM, Langlo KAR, Aksetøy I-LA, Lundgren KM, Skovereng K, Sandbakk Ø, Wisløff U, Dalen H. Indexing cardiac volumes for peak oxygen uptake to improve differentiation of physiological and pathological remodeling: from elite athletes to heart failure patients. Eur. Heart J. Cardiovasc. Imaging. 2023;24:721–729.

30. Bletsa E, Oikonomou E, Dimitriadis K, Stampouloglou PK, Fragoulis C, Lontou SP, Korakas E, Beneki E, Kalogeras K, Lambadiari V, et al. Exercise Effects on Left Ventricular Remodeling in Patients with Cardiometabolic Risk Factors. Life. 2023;13:1742.

31. Nayor M, Xanthakis V, Tanguay M, Blodgett JB, Shah RV, Schoenike M, Sbarbaro J, Farrell R, Malhotra R, Houstis NE, et al. Clinical and Hemodynamic Associations and Prognostic Implications of Ventilatory Efficiency in Patients With Preserved Left Ventricular Systolic Function. Circ. Heart Fail. 2020;13:e006729.

32. Lauer MS, Larson MG, Evans JC, Levy D. Association of left ventricular dilatation and hypertrophy with chronotropic incompetence in the Framingham Heart Study. Am. Heart J. 1999;137:903–909.

33. Alexeeff SE, Deosaransingh K, Van Den Eeden S, Schwartz J, Liao NS, Sidney S. Association of Long-term Exposure to Particulate Air Pollution With Cardiovascular Events in California. *JAMA Netw*. Open. 2023;6:e230561.

34. Garshick E, Blanc PD. Military deployment-related respiratory problems: an update. Curr. Opin. Pulm. Med. 2023;29:83–89.

35. Long-Term Health Consequences of Exposure to Burn Pits in Iraq and Afghanistan. Mil. Med. 2015;180:601–603.

36. Masiol M, Mallon T, Haines KM, Utell MJ, Hopke PK. Airborne Dioxins, Furans and Polycyclic Aromatic Hydrocarbons Exposure to Military Personnel in Iraq. J. Occup. Environ. Med. Am. Coll. Occup. Environ. Med. 2016;58:S22–S30.

37. Kutikhin AG, Sinitsky MYu, Yuzhalin AE, Velikanova EA. Shear stress: An essential driver of endothelial progenitor cells. J. Mol. Cell. Cardiol. 2018;118:46–69.

38. Abplanalp W, DeJarnett N, Riggs DW, Conklin DJ, McCracken JP, Srivastava S, Xie Z, Rai S, Bhatnagar A, O’Toole TE. Benzene exposure is associated with cardiovascular disease risk. PLOS ONE. 2017;12:e0183602.

39. Zelko IN, Dassanayaka S, Malovichko MV, Howard CM, Garrett LF, Uchida S, Brittian KR, Conklin DJ, Jones SP, Srivastava S. Chronic Benzene Exposure Aggravates Pressure Overload-Induced Cardiac Dysfunction. Toxicol. Sci. Off. J. Soc. Toxicol. 2021;185:64–76.

40. Lind PM, Orberg J, Edlund U-B, Sjöblom L, Lind L. The dioxin-like pollutant PCB 126 (3,3’,4,4’,5-pentachlorobiphenyl) affects risk factors for cardiovascular disease in female rats. Toxicol. Lett. 2004;150:293–299.

41. Jokinen MP, Walker NJ, Brix AE, Sells DM, Haseman JK, Nyska A. Increase in cardiovascular pathology in female Sprague-Dawley rats following chronic treatment with 2,3,7,8-tetrachlorodibenzo-p-dioxin and 3,3’,4,4’,5-pentachlorobiphenyl. Cardiovasc. Toxicol. 2003;3:299– 310.

42. Humblet O, Birnbaum L, Rimm E, Mittleman MA, Hauser R. Dioxins and Cardiovascular Disease Mortality. Environ. Health Perspect. 2008;116:1443.

43. Marris CR, Kompella SN, Miller MR, Incardona JP, Brette F, Hancox JC, Sørhus E, Shiels HA. Polyaromatic hydrocarbons in pollution: a heart-breaking matter. J. Physiol. 2020;598:227–247.

44. Biswas G, Srinivasan S, Anandatheerthavarada HK, Avadhani NG. Dioxin-mediated tumor progression through activation of mitochondria-to-nucleus stress signaling. Proc. Natl. Acad. Sci. U. S. A. 2008;105:186–191.

45. Kim YH, Warren SH, Kooter I, Williams WC, George IJ, Vance SA, Hays MD, Higuchi MA, Gavett SH, DeMarini DM, et al. Chemistry, lung toxicity and mutagenicity of burn pit smoke-related particulate matter. Part. Fibre Toxicol. 2021;18:45.

46. Marfella Raffaele, Prattichizzo Francesco, Sardu Celestino, Fulgenzi Gianluca, Graciotti Laura, Spadoni Tatiana, D’Onofrio Nunzia, Scisciola Lucia, La Grotta Rosalba, Frigé Chiara, et al. Microplastics and Nanoplastics in Atheromas and Cardiovascular Events. N. Engl. J. Med. 2024;390:900–910.

47. Aung N, Sanghvi MM, Zemrak F, Lee AM, Cooper JA, Paiva JM, Thomson RJ, Fung K, Khanji MY, Lukaschuk E, et al. Association Between Ambient Air Pollution and Cardiac Morpho-Functional Phenotypes. Circulation. 2018;138:2175–2186.

48. Pope CA, Brook RD, Burnett RT, Dockery DW. How is cardiovascular disease mortality risk affected by duration and intensity of fine particulate matter exposure? An integration of the epidemiologic evidence. Air Qual. Atmosphere Health. 2011;4:5–14.

49. Gestro M, Condemi V, Bardi L, Tomaino L, Roveda E, Bruschetta A, Solimene U, Esposito F. Short-term air pollution exposure is a risk factor for acute coronary syndromes in an urban area with low annual pollution rates: Results from a retrospective observational study (2011–2015). Arch. Cardiovasc. Dis. 2020;113:308–320.

50. Falvo MJ, Osinubi OY, Sotolongo AM, Helmer DA. Airborne Hazards Exposure and Respiratory Health of Iraq and Afghanistan Veterans. Epidemiol. Rev. 2015;37:116–130.

51. Engelbrecht JP, McDonald EV, Gillies JA, “Jay” Jayanty RKM, Casuccio G, Gertler AW. Characterizing Mineral Dusts and Other Aerosols from the Middle East—Part 2: Grab Samples and Re-Suspensions. Inhal. Toxicol. 2009;21:327–336.

52. Wenger J, O’Connell C, Cottrell L. Examination of Recent Deployment Experience Across the Services and Components [Internet]. RAND Corporation; 2018 [cited 2024 Jul 29]. Available from: https://www.rand.org/pubs/research_reports/RR1928.html

53. McFarlane AC. The long-term costs of traumatic stress: intertwined physical and psychological consequences. World Psychiatry. 2010;9:3.

54. DeFlorio-Barker S, Lobdelle DT, Stone SL, Boehmer T, Rappazzo KM. Acute effects of short-term exposure to air pollution while being physically active, the potential for modification: A review of the literature. Prev. Med. 2020;139:106195.

55. Zell-Baran LM, Meehan R, Wolff J, Strand M, Krefft SD, Gottschall EB, Macedonia TV, Gross JE, Sanders OL, Pepper GC, et al. Military Occupational Specialty Codes: Utility in Predicting Inhalation Exposures in Post-9/11 Deployers. J. Occup. Environ. Med. 2019;61:1036–1040.

56. Howard JT, Stewart IJ, Kolaja CA, Sosnov JA, Rull RP, Torres I, Janak JC, Walker LE, Trone DW, Armenta RF. Hypertension in military veterans is associated with combat exposure and combat injury. J. Hypertens. 2020;38:1293–1301.

57. Cai Y, Zhang B, Ke W, Feng B, Lin H, Xiao J, Zeng W, Li X, Tao J, Yang Z, et al. Associations of Short-Term and Long-Term Exposure to Ambient Air Pollutants With Hypertension. Hypertension. 2016;68:62–70.

58. Cheng S, Fernandes VRS, Bluemke DA, McClelland RL, Kronmal RA, Lima JAC. Age-Related Left Ventricular Remodeling and Associated Risk for Cardiovascular Outcomes. Circ. Cardiovasc. Imaging. 2009;2:191–198.

